# Emergency dementia crisis care: Exploring health care staff views on crisis care optimisation across emergency services in England

**DOI:** 10.64898/2026.06.08.26355155

**Authors:** Elena Mirea Conley, Georgia Bell, Julia Fountain, Katherine Sykes, Dorina Cadar, Naji Tabet, Alessandro Bosco

## Abstract

**Background:** In the UK, over 36 million contacts are made annually by people living with dementia (PLWD) to either primary or secondary community mental health services. As dementia progresses, PLWD may experience increased distress and resort to 999 calls for an ambulance, which may in turn result in conveyance to Accident & Emergency (A&E). Nearly 1 million A&E attendances are made by PLWD. This trend is set to rise sharply as the prevalence rates of dementia increase over time and as the condition progresses, with associated healthcare costs impacting overall care delivery. This may lead to reduced resource allocation for dementia emergency services, negatively affecting the experiences of both providers and service users.

**Aim(s):** To explore ways to improve access and quality of care to emergency crisis care for PLWD from the perspective of healthcare staff providing this type of support.

**Methods:** This qualitative study explored (1) the experiences, resources, and needs of healthcare professionals in emergency and community settings to support access for PLWD, and (2) the mechanisms influencing dementia crisis response. The COREQ Checklist was used to improve transparency, credibility, and reproducibility. Inter-rater reliability was calculated. PPIE contributors co-developed recommendations for healthcare professionals, and study findings informed a comic-based dissemination resource shared with third-sector organisations (e.g. Alzheimer’s Society) to support community awareness and engagement.

**Results:** Fifteen interviews were held with emergency services staff. Inter-rater reliability was substantial between two raters (κ = 0.62). Four overarching themes, with associated subthemes, were identified relating to crisis care delivery, barriers to effective response, and strategies employed to address these challenges. Additional themes captured decision-making processes at key points in the care pathway, including initial crisis response, during intervention, and at discharge from emergency and community services. Decision-making was characterised by the need to balance patient safety with autonomy in determining care in the best interests of PLWD and their informal carers.

**Discussion:** This exploratory study reveals frontline staff perspectives on challenges and actionable strategies for dementia crisis care. Findings support targeted service improvements, cross-sector collaboration, and co-produced resources to enhance outcomes for PLWD and their informal carers.

## Background

People living with dementia (PLWD) frequently access urgent and emergency care services during episodes of acute behavioural disturbance or psychological distress. Emergency ambulance callouts and subsequent conveyance to acute hospitals are common responses to such crises, with studies showing high rates of ambulance utilisation and hospital transport among older adults with dementia and multimorbidity (Voss et al., 2017; Han et al., 2024). However, unplanned hospital admissions can exacerbate cognitive disorientation and contribute to physical deconditioning and are associated with poorer outcomes for this population (Voss et al., 2017; Alzheimer’s Society, 2024). These adverse outcomes are linked to prolonged inpatient stays and delayed discharge, which in turn place pressure on emergency departments and ambulance services. Evidence indicates that people with dementia experience significantly longer lengths of stay and increased risk of mortality following hospital admission compared with those without dementia (Reeves et al., 2023; Di Martino et al., 2025).

In the United Kingdom (UK), PLWD represent a substantial proportion of emergency care demand, accounting for nearly one million Accident & Emergency (A&E) attendances annually (Alzheimer’s Society, 2024). Healthcare utilisation is disproportionately high across dementia populations, particularly among individuals with undiagnosed or early-stage disease, who are significantly more likely to attend emergency departments (Alzheimer’s Society, 2024). Compared with patients without dementia, PLWD are more likely to experience longer hospital stays following unplanned admissions, and those in advanced stages of the condition are at greater risk of prolonged hospitalisation and adverse outcomes, including mortality (Reeves et al., 2023; Di Martino et al., 2025).

Although integrated neighbourhood teams and other community-based care models are being developed to enhance coordination across primary, community, mental health, and voluntary services, the emergency care pathway for dementia remains poorly defined (Department of Health and Social Care, 2026). Existing evidence suggests considerable variation in crisis service provision and care pathways, as well as limited availability of alternative community responses to avoid hospital admission (Streater et al., 2017; Han et al., 2024). Strengthening the capacity of community-based services and clarifying crisis response pathways are therefore critical to reducing avoidable emergency attendances and hospital admissions. A systematic examination of the resources, barriers, and decision-making processes encountered by healthcare professionals across ambulance services, acute care, geriatrics, and community crisis teams is needed to inform interventions that optimise crisis management and improve outcomes for PLWD and their informal carers (Han et al., 2024; Voss et al., 2017).

## Methods

### Study design

This was a qualitative exploratory study utilising semi-structured interviews with health care professionals involved in the delivery of dementia emergency and/or crisis service responses. We used a socio-constructivist framework (Berger & Luckmann, 1967) and described dementia crisis care as a complex adaptive system, in which ‘crisis’ emerges as a socially situated and system-mediated process rather than a discrete clinical episode.

### Recruitment and data collection

The study protocol was developed in collaboration with the Alzheimer’s Society volunteer network, the ARC KSS implementation manager, and academic partners. Key stakeholders involved in dementia service provision across Kent, Surrey and Sussex were identified through targeted web searches and existing professional networks at the Centre for Dementia Studies at Brighton and Sussex Medical School, the University of Sussex, the University of Brighton, and ARC-KSS. Additional participants were recruited using snowball sampling. Potential participants were initially contacted by email with a brief study summary; those who expressed interest were sent an information sheet and consent form and offered a remote interview at a mutually convenient time.

Supplementary recruitment was facilitated via advertisements and partner organisations, including the Brighton Medico-Chirurgical Society, Dementia UK, the Integrated Academic Training programme at Brighton and Sussex Medical School, and the Kent, Surrey and Sussex Regional Delivery Network ageing speciality research network. Interested individuals responded directly to the researcher by email or telephone. All participants were invited to take part in a semi-structured interview of approximately 60 minutes. Interviews were scheduled to suit participants’ availability and conducted remotely (by telephone or secure videoconference) according to participant preference. With participant consent, interviews were audio-recorded using a password-protected digital recorder; where recording was not possible, contemporaneous notes were taken. Participants who completed an interview received a £30 voucher as reimbursement for their time. An exemplar interview guide is provided in Appendix 1.

Interview questions were tailored to the participant’s service context (ambulance services; emergency department/acute inpatient care; community crisis and integrated response services) and broadly explored: Key challenges and barriers to providing effective dementia support in the community; policy and planning considerations, including the extent to which dementia needs are reflected in strategy and service delivery; required infrastructures to optimise crisis response for PLWD; examples of good practice and opportunities for service improvement.

#### Patient and Public Involvement and Engagement

A Patient and Public Involvement and Engagement (PPIE) meeting was held in April 2026, co-delivered with a PPIE facilitator. Ten members participated, and a professional artist produced visual vignettes based on the preliminary analysis. This meeting also served as a bridge between the qualitative findings and the development of a grant proposal on the same topic. These procedures were designed to enhance analytic rigour, transparency, and stakeholder validity, while recognising the inherently reflexive nature of qualitative interpretation.

### Reflexivity

To mitigate researcher effects during interviews, the fellow used open-ended prompts, neutral and non-judgmental language, and a consistent topic guide tailored to each service context. Interview techniques emphasised active listening and clarification rather than leading questioning. During analysis, multiple researchers independently coded a subset of transcripts and compared coding decisions to enhance analytic transparency. Patient and public involvement contributors reviewed emergent themes to ensure interpretations resonated with lived experience and to reduce the risk of investigator bias. Collectively, these procedural safeguards aimed to increase the credibility and trustworthiness of the findings by making the researcher’s positionality explicit, by documenting its potential influence, and by incorporating triangulation and stakeholder validation into the analytic process.

### Data analysis

Interview transcripts and fieldnotes were managed and organised in NVivo 15 © to facilitate systematic coding and retrieval. Analysis followed the six-phase thematic analysis framework described by Braun and Clarke (2021). Two researchers independently familiarised themselves with the data and generated initial codes; coding was then iteratively reviewed and refined through regular analytic meetings to develop candidate themes and subthemes aligned with the study objectives.

Coding decisions and theme development were documented to maintain an audit trail. Analysis proceeded concurrently with data collection to allow emergent findings to inform subsequent topic-guide refinements and sampling decisions. A codebook was developed to guide the systematic coding of the transcripts. To assess inter-rater reliability, one full transcript was independently coded by two researchers, and Cohen’s kappa coefficient was calculated. Following this, all transcripts were fully coded, and digital tools were used to support organisation and visualisation of coded data, while interpretation and theme development remained researcher led.

## Results

A total of 15 semi-structured qualitative interviews were conducted with healthcare professionals from a range of emergency and community-based services, including ambulance staff (n=4), Admiral and mental health nurses (n=4), consultant geriatricians (n=3), dementia link workers (n=3), and a head of operations in independent living provision for older people with cognitive impairment (n=1). Analysis of interviews with healthcare professionals across acute, community, and emergency settings identified four interrelated themes and associated subthemes that describe how dementia crisis care is experienced and delivered. These themes capture system-level constraints, relational dynamics, and adaptive practices, highlighting how crisis emerges not as a discrete clinical event but as a system-mediated and socially situated phenomenon (Table 1 for coded texts across transcripts). To enhance analytic rigour and transparency, a subset of transcripts was independently reviewed by two researchers to support discussion and refinement of the coding framework. Coding and theme development were discussed iteratively within the research team throughout (κ =0.62).

**Table 1.** Reporting of most significant coded text across transcripts.

| **Themes** | **Code** | **Example Quote** | **N codes across transcripts** |
| --- | --- | --- | --- |
| 1. Problems affecting crisis care delivery | **1a. Signposting (unclear routes)** | “families... don’t have a clue what we’re doing or how we can support... they got the diagnosis and then it’s like... what now (Participant 11, Admiral nurse) | 27 |
|  | **1b. Fragmented records and access** | “during hospital stay... difficult to get information... staff asked us for information rather than giving us information... caused frustration...” (Participant 13, Head of Operations) | 21 |
| 2. Readiness to response and unmet needs | **2a. Strategies in place to deal with dementia crisis** | “Inside the trust we do have... Dementia and Delirium Team... specialist nurses... help manage anybody who's an inpatient... either around a new diagnosis or somebody who's come in... in crisis...” (Participant 14, Consultant geriatrician) | 65 |
|  | **2b. Limited psychological support for staff** | “It is and one of the things that we have in place for the care teams They also have the opportunity to talk about how it feels, supporting someone in real distress...It’s so important that they get the opportunity to talk about how they feel, so that they are actually able to keep giving rather than become very burnt out.” (Participant 15, Admiral nurse) | 17 |
|  | **2c. Distress as unmet needs (non-medical vs systemic)** | “Urgency is because everybody is just confused... the systems don’t work well... GP, hospital, outreach... everyone works in their.. little bubble... so that is why situations become urgent,... nobody is really linking things together.” (Participant 19, Admiral nurse) | 53 |
| 3. Crisis care decision making | **3a. Family as essential narrators and decision initiators** | “if someone has capacity they go home... if they lack capacity... best interest decisions... involving family where possible...” (Participant 16, Admiral nurse) | 49 |
|  | **3b. Professionals’ orientation to autonomy vs family safety concerns** | “They might refuse everything... informal carers are overwhelmed, trying to put support in place but the person won’t accept anything... and then it becomes about whether to overrule or not... sometimes you have to take responsibility and do what’s right.” (Participant 18, Dementia Link worker) | 24 |
| 4. Discharge & Follow-up | **4a. Follow-up care / readmission / continuity** | “We don’t follow up, ever... once we’ve dropped a patient off at hospital that’s us done... unless we happen to see them again, we’ve got nothing to do with them... so there’s no real continuity after that point.” (Participant 20, Ambulance staff) | 41 |

### 1. Problems affecting crisis care delivery

Participants consistently described unclear and fragmented service pathways, with limited coherence across health and social care systems. Signposting processes were frequently experienced as confusing, inconsistent, and burdensome for patients and informal carers. Rather than facilitating access, service navigation was described as requiring substantial effort from families, who were often expected to identify and engage appropriate services independently.

> *‘The systems don’t work well… everyone kind of works in their own little bubble… people get multiple phone calls… is it the Council, is it the NHS? Nobody seems to tell me exactly what I can get… so in the end they just say, I’d rather manage myself*.*’ (Participant 19, Admiral nurse)*

This lack of clarity was particularly problematic in the context of dementia, where cognitive impairment limited the ability of patients to engage with complex systems. Participants suggested that unclear signposting contributed to delays in care and increased the likelihood of crisis escalation. Participants reported significant challenges related to limited access to patient information across organisational boundaries. Records were often incomplete, inaccessible, fragmented or inconsistent across systems, leading to uncertainty in clinical decision-making.

Ambulance and community-based professionals highlighted difficulties accessing up-to-date information, as records had transitioned to digital platforms not universally available across services:

> *‘…And because all of the care notes are now online, she has care. It’s four times a day, I think. But we can’t access any of their records, which is something that’s quite unhelpful these days because it always used to be on paper…’ (Participant 20, Ambulance staff)*

These informational gaps led to risk-averse practices, including hospital admission without clear clinical need, due to concerns about patient safety. Participants described this as a systemic issue in which no single professional or service had a complete picture of the patient’s needs, resulting in episodic and fragmented care delivery.

### 2. Readiness to response and unmet needs

Participants highlighted practical strategies and the emotional demands associated with providing dementia crisis care, including managing distress, supporting families, and making complex decisions under uncertainty. Practical strategies for managing crisis situations were often implemented in response to system limitations. These strategies included: temporarily “holding” patients in acute settings, extending time spent with patients and families, coordinating across multiple services, prioritising immediate stabilisation over long-term resolution:

> *‘Or contact the crisis team if that’s what I feel is most appropriate, but we’ll try and manage it. So that I mean, part of my job is to keep people out of hospital, so we’ll manage his best we can*…*’ (Participant 21, Dementia Link worker)*

These approaches were often informal and dependent on individual professional judgement, rather than guided by structured pathways. Participants described taking on coordination roles beyond their formal remit in order to manage complexity. Access to structured psychological support was reported to be limited. Support, where available, was often informal and inconsistently applied.

> *“We don’t get much support… we have a weekly meeting where we discuss difficult cases… distressed patients, family conflict… but it’s quite informal*.*’ (Participant 19, Admiral nurse)*

The absence of structured support mechanisms meant that staff often managed emotional demands independently or through peer support. This raised concerns about sustainability, particularly given repeated exposure to complex, unresolved crisis situations. Participants consistently framed the crisis in dementia as arising from unmet needs, rather than solely from acute medical deterioration:

> *‘Generally, urgency is because everybody is just confused and this the systems don’t work well the…Different services don’t kind of link up the GP to the hospital, to the outreach to the everyone kind of works in their own little bubble. So that’s why it’s… urgent*.*’ (Participant 19, Admiral nurse)*

In this context, a crisis was often precipitated by accumulated system pressures, with non-medical needs becoming medicalised due to a lack of alternative responses.

### 3. Crisis care decision making

This integrated theme highlights that decision-making in dementia crisis care is not solely a clinical task, but a socially mediated process shaped by relationships, system constraints, and ethical ambiguity. Decision-making in dementia crisis care was described as a fundamentally relational and context-dependent process, shaped by the interplay between family involvement, patient capacity, and professional responsibility for managing risk. Participants consistently positioned families, particularly informal carers, as central to decision-making processes, serving as primary sources of information, initiators of care, and coordinators across fragmented services. Informal carers were often responsible for recognising early signs of deterioration, communicating patient history, and facilitating engagement with services. In the absence of such support, decision-making processes were described as significantly compromised:

> *‘If there’s a carer on scene, we will always eventually…speak to them about their point of view…But generally, our first approach would be to speak to the patient…if there’s no informal carers around, we… find a next of kin number…*.*with their permission, have a Snoop around the house and look for discharge summaries or prescription lists*…*’ (Participant 20, Ambulance staff)*

At the same time, reliance on families introduced variability and additional complexity. Carer capacity, levels of understanding, and emotional burden influenced both the timing and nature of decision-making. In some cases, informal carers experiencing high levels of stress or confusion struggled to engage with services or make informed decisions about care:

> *‘Obviously part of that work means that almost on a daily basis, some of the folk that I’m supporting will go into crisis… it may be crisis where the carer becomes ill… or the person living with dementia falls off a cliff…or they’re in great anxiety and fear… so quite often I will go out… and I’ll sit with the family… until I can get a handle on what’s going wrong…’ (Participant 21, Dementia link worker)*

Within this relational context, professionals described navigating ongoing ethical tensions, particularly the balance between respect for patient autonomy and concerns about safety and risk. These tensions were often heightened in situations involving uncertainty about mental capacity, differing perspectives between patients and families, and safeguarding concerns:

> *‘Sometimes you have situations where safeguarding might need to be considered… but actually a lot of the time it’s about the relationships and what’s going on between family members… You end up working with that as much as the patient*.*’ (Participant 16, Admiral/mental health nurse)*

Decision-making was therefore not experienced as a linear or protocol-driven process but rather as a negotiated practice that required continuous assessment of risk, capacity, and relational dynamics. Professionals frequently described making situational judgements under conditions of uncertainty, and in some cases, withdrawing and seeking additional support when risk escalated. Participants described navigating complex ethical tensions, particularly in balancing patient autonomy with concerns regarding safety:

> *‘Sometimes you’re going into situations where you’re not entirely sure what you’re going to find… You have to make a judgement there and then… and if it feels too risky, you step back and get other services involved*.*’ (Participant 12, Dementia Adviser)*

Decision-making was described as context-dependent and negotiated, rather than guided by clear protocols. Professionals often had to make judgments under conditions of uncertainty, balancing competing priorities.

### 4. Follow-up care

Across all transcripts, follow-up care emerged as a critical point of system weakness, characterised by inconsistency, fragmentation, and limited capacity to address ongoing needs. Participants described how insufficient or poorly coordinated follow-up contributes directly to repeated crises and cycling through services. Rather than representing a discrete phase of care, follow-up was experienced as variable, contingent, and often inadequate, leaving patients in a state of *precarious stability*. Participants across settings reported that care provided during or immediately after a crisis was often not sustained over time, with limited mechanisms to ensure continuity:

> *‘Quite often we’re taking them into hospital just because we have no idea what’s going on… we can’t safely leave them at home when we’ve got absolutely no clue…’ (Participant 20, Ambulance staff)*

Follow-up was therefore not consistently embedded within care pathways, leading to situations where: underlying needs remained unaddressed, deterioration progressed unchecked, and crisis became the point of re-entry into the system.

## Discussion

Overall, the results demonstrate that dementia crisis care operates within a constrained and fragmented system, requiring continuous adaptation by both professionals and families. Crisis is best understood not as an isolated event but as the outcome of interacting system-level, relational, and contextual factors that together shape patterns of repeated service use and ongoing unmet need. This study demonstrates that decision-making in dementia crisis care is relational, distributed, and context-dependent, shaped by interactions among family involvement, patient capacity, and professional responsibility for managing risk. In parallel, findings highlight that inadequate follow-up and continuity of care are central drivers of recurrent crises, contributing to repeated use of emergency services. Together, these findings suggest that dementia crisis care is not best conceptualised as a series of discrete clinical events, but as a system-mediated trajectory, in which decision-making and outcomes are contingent on relational networks and continuity of care. The centrality of families in decision-making is strongly supported by recent literature. A systematic review of emergency department care for PLWD found that care partners play a critical role in providing clinical information, supporting decision-making, and coordinating care, particularly where communication and system integration are limited (Jelinski et al., 2025). Similarly, studies of decision-making in advanced dementia highlight that, as cognitive capacity declines, informal carers increasingly assume responsibility for making complex health decisions, often in collaboration with professionals (Davies et al., 2022).

The present findings extend this literature by demonstrating that family involvement is not merely supportive but structurally necessary for effective decision-making in crisis contexts. Participants described how, in the absence of a carer, decision-making processes become significantly constrained, reflecting existing research on the reliance on “care partners” to bridge communication and information gaps in emergency settings (Jelinski et al., 2025). However, consistent with broader dementia research, this study also highlights the burden and variability associated with caregiving. High levels of stress, emotional exhaustion, and uncertainty among informal carers are widely reported, with evidence indicating that caregiver burden can significantly affect decision-making capacity and engagement with services (Jeste et al., 2021). This aligns with current findings, in which overwhelmed informal carers were less able to engage with services or make informed decisions, contributing to delays and fragmentation in care (Jeste et al., 2021).

The study’s finding that professionals engage in situated, negotiated decision-making under conditions of uncertainty is also supported by emerging evidence from emergency and pre-hospital care settings. Qualitative studies of ambulance clinicians show that decision-making for people with dementia is influenced not only by clinical factors but also by cognitive capacity, patient circumstances, and available social support, with professionals often relying on incomplete information and contextual judgement (Voss et al., 2020). More recent work further emphasises the ethical dimension of this process. Research indicates that clinicians frequently experience tension between respecting autonomy and ensuring safety, particularly when patients have impaired decision-making capacity, leading to reliance on substitute decision-making and negotiation with families. These findings closely mirror those of the present study, in which participants described balancing competing priorities of autonomy, risk, and feasibility in real time (Bennesved et al. 2023). Importantly, the current analysis advances this understanding by highlighting how these ethical tensions are amplified by system constraints, including fragmented information and limited-service options. Decision-making is therefore shaped not only by clinical and ethical considerations but also by structural limitations, reinforcing the need to conceptualise it as a system-level phenomenon. The role of follow-up care in driving recurrent crises is strongly supported by recent evidence. Qualitative research examining emergency department transitions for PLWD found that poorly coordinated discharge processes and inadequate follow-up care are key contributors to adverse outcomes and repeated service use (McHugh et al., 2024). Quantitative evidence similarly demonstrates that continuity of care is associated with improved outcomes. For example, population-based studies indicate that higher continuity of primary care is associated with reduced hospital admissions in people with dementia (Leniz et al., 2022), suggesting that sustained and coordinated care can prevent escalation to crisis.

Conversely, limited follow-up after emergency department discharge is common, with substantial proportions of patients experiencing revisits and hospitalisation within 30 days (Lin et al., 2022). The present findings extend this evidence by showing how inadequate follow-up contributes not only to adverse outcomes but to the production of cyclical crisis trajectories. Participants described patients as existing in a state of “precarious stability,” in which unresolved needs persist until re-escalation occurs. This supports the view that the crisis in dementia is not episodic but recurrent, driven by ongoing gaps in continuity and coordination. The findings support a conceptualisation of dementia crisis care as a complex, adaptive system, characterised by: Relational decision-making, distributed across families and professionals; ethical uncertainty, particularly in balancing autonomy and safety; structural fragmentation, limiting access to information and services; temporal discontinuity, with insufficient follow-up, sustaining cycles of crisis.

Within this system, families operate as a form of informal infrastructure, often compensating for gaps in service coordination and continuity. Professionals, in turn, engage in adaptive and negotiated practices to manage clinical and social risk in real time. These findings indicate that dementia crisis care is shaped by both relational dynamics and system-level constraints, with decision-making distributed across families and professionals under conditions of uncertainty. Limited follow-up and poor continuity of care were consistently reported, contributing to recurrent cycles of crisis and instability. Collectively, this underscores the need for more integrated, sustained, and person-centred models of care that better align with the complex and evolving needs of people living with dementia and their informal carers.

### Strengths and limitations

This study addresses an important and underexplored aspect of dementia care by examining crisis response pathways across emergency and community healthcare services in England. Given the increasing prevalence of dementia and growing pressures on urgent and emergency care systems, understanding how crisis care is experienced and delivered is of considerable clinical and policy relevance. A key strength of the study is the inclusion of a multidisciplinary sample of healthcare professionals working across ambulance services, geriatrics, dementia support services, mental health nursing, and community care. This enabled exploration of dementia crisis care from multiple professional perspectives and facilitated a more comprehensive understanding of the organisational and relational complexities of crisis response. The qualitative design enabled an in-depth exploration of frontline experiences, ethical tensions, and system-level barriers that are often difficult to capture with quantitative approaches alone. Rich participant narratives provided detailed insight into how healthcare professionals navigate uncertainty, fragmented systems, and competing priorities when supporting people living with dementia and their informal carers. Another strength of the study is its conceptual framing of dementia crisis care as a relational and system-mediated process rather than a discrete clinical event. This systems-oriented perspective contributes to the growing literature emphasising continuity of care, integrated service delivery, and the importance of relational decision-making in dementia care pathways. The study also benefited from meaningful PPIE. PPIE contributors were involved in developing recommendations and dissemination materials, helping ensure that the findings remained grounded in lived experience and relevant to practice and service development. Methodological rigour was further enhanced through the use of the COREQ checklist, iterative coding discussions, maintenance of an audit trail, and collaborative theme development among researchers. These processes strengthened transparency and credibility.

Several limitations should be acknowledged. First, the study involved a relatively small sample of healthcare professionals recruited primarily from Kent, Surrey, and Sussex. Although qualitative research does not seek statistical generalisability, the transferability of findings to other geographical regions or healthcare systems may therefore be limited. Second, the study focused exclusively on professional perspectives and did not directly include the experiences of PLWD or informal carers. While healthcare staff provided valuable insights into crisis care delivery, future research incorporating lived-experience perspectives would yield a more comprehensive understanding of dementia crisis pathways. The study may also be subject to self-selection bias, as participants with a particular interest or experience in dementia care may have been more likely to participate. Consequently, the findings may overrepresent the perspectives of highly engaged professionals. Most interviews were conducted remotely via telephone or videoconferencing, which may have limited opportunities to observe non-verbal communication and contextual factors that can enrich qualitative data collection. Although the study included participants from several healthcare settings, some professional groups involved in dementia crisis care, including general practitioners, emergency department physicians, social workers, and commissioners, were underrepresented or absent. Their inclusion may have provided additional perspectives on system coordination and decision-making. Finally, although the study adopted a thematic analysis approach, the inclusion of inter-rater reliability statistics may be viewed as methodologically inconsistent by some qualitative researchers, given that thematic analysis does not typically prioritise coder agreement as a marker of analytic rigour. This study suggests that dementia crisis care is best understood not as an isolated emergency event, but as the consequence of cumulative fragmentation across health, social, and relational systems. Improving outcomes for PLWD will require integrated and sustained models of care that support continuity, strengthen coordination across sectors, and recognise informal carers as central partners in crisis prevention and response.

## Data Availability

All data (anonymised) produced in the present study are available upon reasonable request to the authors

