## Appendix 1 for "Emergency dementia crisis care: Exploring health care staff views on crisis care optimisation across emergency services in England"

**Appendix 1. Topic guide: Interviews with healthcare professionals.**

### Semi-structured topic guide

Title: Emergency dementia crisis care: Exploring health care staff views on crisis care optimisation across emergency services in England.

Questions will be reshaped according to data from previous interviews and as suggested by our partnered PPI group.

#### Instructions:

1. The interview lasts around 60-minutes.
2. Make introduction at the start of each session. This will take around 5 minutes.
3. After the interview, please write a short (2-3 paragraph) reflective note.
4. Audio record the session.
5. Use one voice recorder.

#### Introductions (5 minutes)

1) **The research fellow introduces themselves and the project.**
2) **The research fellow explains about taking part in the interview:**

#### Semi-structured interviews (55 minutes):

**Introduction (5 minutes):**

Ask participants to introduce themselves by giving their name, job title and how long they have been involved in supporting people in emergency/crisis care in dementia.

**Topic 1. How cases are assessed (15 minutes):**

1. What type of support do you usually provide to patients about emergency (or crisis depending on role) support?
   - *Ask for examples to illustrate points made – could you explain a little more about what that has looked like in practice?*
2. Some patients may come to you in a crisis. How do you assess which support is most relevant for them?
   - *Ask for examples to illustrate points made – could you explain a little more about what that has looked like in practice?*
3. What are the characteristics of crisis experience (or distressed behaviour) ?
   - *Ask for examples to illustrate points made – could you explain a little more about what that has looked like in practice?*

**Topic 2. What barriers are present in the process of providing support (15 minutes):**

4. What barriers do you encounter when you support patients and their families?
   - *Ask for examples to illustrate points made – could you explain a little more about what has helped them in their support to patients' and families experiences, what has not helped, and why?*

5. What barriers do you encounter when you have to get in contact with other services (this could be community crisis services) ?
    - *Ask for examples to illustrate points made – could you explain a little more about why this has helped or been a hinderance?*

**Topic 3. What enablers support the process of providing support (15 minutes):**

6. What do you feel are the enablers in your work? What is it that you do (part of your support) that is particularly helpful for patients and their family?
    - *Ask for examples to illustrate points made – could you explain a little more about why this has helped or been a hinderance?*

**Topic 4. Looking to the future (15 minutes):**

7. Looking to the future, is there anything about your practicing that you would like to keep?
8. Are there some changes you think should be made to effectively support patients and their families in accessing emergency/crisis support?

**Ending session (5 minutes):**

9. Ask if they have other comments they wish to make.
10. Thank participant for participating. Re-iterate confidentiality.

#### Reflections

Please write a short reflection below:
